# Early School Years follow up of the Asking Questions in Alcohol Longitudinal Study in Melbourne, Australia (*AQUA at 6*): Cohort profile

**DOI:** 10.1101/2021.06.17.21259124

**Authors:** Evelyne Muggli, Jane Halliday, Elizabeth Elliott, Anthony Penington, Deanne K Thompson, Alicia Spittle, Della A. Forster, Sharon Lewis, Stephen Hearps, Peter J Anderson

## Abstract

**Purpose:** The Asking Questions about Alcohol in Pregnancy (AQUA) study, established in 2011, is a pre-birth cohort of 1570 mother and child pairs designed to assess the effects of low to moderate prenatal alcohol exposure and sporadic binge drinking on long-term child development. The current follow-up of the children, now aged 6 to 8 years, aims to strengthen our understanding of the relationship between these levels of prenatal alcohol exposure and neuropsychological functioning, facial dysmorphology, and brain structure & function.

**Findings to date:** Over half (59%) of mothers consumed some alcohol during pregnancy, with one in five reporting at least one binge drinking episode prior to pregnancy recognition. Children’s craniofacial shape was examined at 12 months of age, and low to moderate prenatal alcohol exposure was associated with subtle midface changes. At two years of age, formal developmental assessments showed no evidence that cognitive, language or motor outcome was associated with any of the prenatal alcohol exposures investigated.

**Participants:** Between June 2018 and April 2021, 802 of the 1342 eligible AQUA study families completed a parent-report questionnaire (60%). Restrictions associated with COVID-19 pandemic disrupted recruitment, but early school-age neuropsychological assessments were undertaken with 696 children (52%), and 482 (36%) craniofacial images were collected. A pre-planned, exposure-representative subset of 146 random children completed a brain MRI. The existing AQUA study biobank was extended through collection of 427 (32%) child buccal swabs.

**Future plans:** We will investigate the relationship between prenatal alcohol exposure and specific aspects of neurodevelopment at 6-8 years, including brain structure & function. We will also determine whether craniofacial changes identified at 12 months of age are predictive of later developmental impairments. The contribution of genetics and epigenetics to individual variations in outcomes will be examined in conjunction with established and future national and international collaborations.

**STRENGTHS AND LIMITATIONS:**

- The Asking Questions about Alcohol in Pregnancy (AQUA) cohort study was specifically designed to prospectively collect high-quality data on low to moderate prenatal alcohol exposure and relevant confounders to investigate the risk to offspring neurodevelopment.
- The children are being followed up for the third time at 6-8 years, using sensitive measures of neuropsychological function, 3D craniofacial photography, and brain MRI.
- A biobank of birth samples and maternal and child buccal DNA enables investigation of the contribution of genetic and epigenetic factors to neurodevelopmental outcomes.
- Despite carefully designed questions, reporting bias will need to be considered in the interpretation of findings, especially around alcohol use.
- The generalisability of some findings will be limited to a general antenatal population of Caucasian women, from middle-income backgrounds and with a low-risk pregnancy.

## INTRODUCTION

Alcohol crosses the placenta and is teratogenic.[1, 2] Health guidelines around the world, including those developed by the Australian National Health and Medical Research Council (NHMRC), recommend that women who are either pregnant or planning a pregnancy abstain from drinking alcohol[3]. Alcohol can damage the developing fetal brain through oxidative stress, damage to the mitochondria, and interference with the function of growth factors and neurotransmitters, as well as through epigenetic changes which regulate gene activity.[2, 4, 5] The consequences can be devastating at a foundational stage of brain development, ultimately disrupting neuronal proliferation and migration, and glial functioning.[2]

High levels of alcohol exposure to the fetal brain can cause a spectrum of structural brain abnormalities, facial dysmorphology, neurological problems and neurodevelopmental impairments, collectively termed Fetal Alcohol Spectrum Disorder (FASD).[4, 6, 7] These effects have been replicated in animal models and are undisputed.[2]

Many pregnant women consume some alcohol during pregnancy, especially around the time of conception.[8-11] This is extremely concerning given the potential harms of prenatal alcohol exposure (PAE) to the developing fetus. Unplanned pregnancy is a common explanation for early pregnancy drinking, particularly for binge drinking exposure.[12] However, even after pregnancy awareness a substantial proportion of women continue to drink at low to moderate levels,[8] sometimes with the knowledge that PAE has the potential to lead to lifelong disabilities in a child.[13] The lack of convincing evidence of harm from lower levels of PAE,[14, 15] and conflicting messages from health professionals concerning adverse effects of low to moderate PAE on the fetus are reasons given by some women for their decision not to abstain.[16]

The effects of PAE vary between individuals likely due to genetic, metabolic, nutritional, social, and environmental factors, as well as the timing, duration, and dose of alcohol.[17] Human research has provided limited evidence that low to moderate PAE is detrimental to the offspring, with a recent systematic review reporting adverse effects on early child development in six studies, no effect in five studies, and a weak positive effect in two.[18] The authors concluded that conflicting findings following low PAE may in part be due to a lack of sensitivity for detecting some outcome measures, and inadequate accounting for confounding, environmental and social factors. Since this review was published, a secondary analysis of 9,719 children from the Adolescent Brain Cognitive Development Study found that even children with low PAE demonstrated poorer psychological and behavioural outcomes at around 9-10 years of age. The authors claimed their findings were robust because potential confounding factors were considered, and that stringent demographic matching procedures increased the plausibility of the findings, but while the study’s sample size is impressive, collection of exposure and confounder information occurred retrospectively in pre-adolescence, raising questions around recall and accuracy.

The Asking Questions about Alcohol in Pregnancy (AQUA) prospective cohort study was designed to address the limitations in exposure measurement and collection of confounders, allowing for a robust investigation of the effects of common drinking patterns in pregnancy.[19]

The primary objective of this current follow-up of the cohort (*AQUA at 6*) is to assess neurodevelopment (neuropsychological functioning, brain structure and function and craniofacial shape) in a population-based cohort of children aged 6-8 years with respect to their PAE (none, low, moderate, high or binge level alcohol exposure), taking into account related maternal, child and socio-environmental factors that may explain individual differences in outcome.

### Hypotheses

1. Any PAE has the possibility of being associated with craniofacial changes (e.g. mid-face, nose, lips and eyes), structural brain changes (e.g. corpus callosum, basal ganglia, cerebellum), and subtle neuropsychological deficits (e.g. motor, attention, executive function, memory and behaviour) at 6-8 years of age;
2. These PAE associations will be influenced by the timing and quantity of alcohol exposure, individual child and maternal characteristics (e.g. genetics, nutrition, breastfeeding, maternal mental health), and socio-environmental factors (e.g. education, lifestyle, parenting style);
3. Craniofacial differences at 12 months of age will be associated with outcomes at 6-8 years, specifically a) craniofacial shape, b) brain structure, and c) neuropsychological functioning.

## COHORT DESCRIPTION

The Asking Questions about Alcohol (AQUA) study comprises a cohort of mother/child dyads recruited from the general population in early pregnancy for longitudinal observation. All women with a singleton pregnancy, attending their first antenatal appointment before 19 weeks gestation, between 25 July 2011 and 30 July 2012, at one of seven public hospital recruitment sites in metropolitan Melbourne, Australia, were eligible to participate. Being 16 years or older and being able to read and write English were prerequisites for participation. The methods are described in detail in the original study protocol.[19] During pregnancy, women completed three questionnaires, and after birth, questionnaires were sent at 12 and 24 months to women who had completed the three pregnancy questionnaires, and for whom complete PAE information was available (n=1570). An exposure representative sub-sample of 850 children were sequentially invited to have a 3D craniofacial photo taken at 12 months (517 images taken), and/or a neurodevelopmental assessment using the Bayley Scales of Infant and Toddler Development (Bayley-III) at 24 months (554 assessments completed).

The cohort of children were recruited again aged between 6 and 8 years, for further assessments, including longitudinal 3D analysis of craniofacial shape, state-of-the-art neuroimaging, and standardised neuropsychological measures to assess neurodevelopmental status. Outcome measure details are provided in a dedicated section below.

### Study design and procedures

Of the 1570 mother and child dyads from the original cohort, 55 mothers had withdrawn from the study. We excluded 108 who were lifetime alcohol abstainers because our target population was children of mothers who normally drink some alcohol. Another 59 mothers were excluded who could not be classified because they abstained in the first trimester, then averaged an intake of less than one standard drink per week for the remainder of their pregnancy. [8]. Therefore, in the *AQUA at 6* follow-up study, 1348 mothers and children were invited to participate. Following the invitation to take part, a further six families were excluded from *AQUA at 6*, because of a recent oncology diagnosis in the child (n=3) or because of a later diagnosed condition impacting long-term development (one child with Down syndrome, one child with Dopa Responsive Dystonia and another child with Sanfilippo Syndrome). The final number of families eligible to participate was 1342.

Data were collected between June 2018 and April 2021.

Neuropsychological assessments and 3D craniofacial imaging were performed in specialist facilities at the Murdoch Children’s Research Institute and Royal Children’s Hospital in Melbourne, Australia (RCH). For families unable to travel to the campus the neuropsychological assessments were administered in the home, at school or another suitable facility such as a library meeting room. Externally assessed test results were obtained when the child had been recently assessed. A PAE-representative subset of children were sequentially invited to have a brain MRI scan, with a target number of 50 in each of three exposure groups: (1) no PAE; (2) PAE in trimester one only *and*; (3) PAE throughout gestation. Primary caregivers (i.e., the AQUA study mother in most cases) completed questionnaires online. This questionnaire was also offered to families whose child did not attend a neuropsychological assessment, but who still wished to take part.

For study participation, the neuropsychological assessment and/or questionnaire needed to be completed. All other aspects of the study were optional.

### Impact of Covid-19

Following the COVID-19 pandemic, adaptations to the assessment procedures were necessary to comply with relevant institutional and government guidelines for a safe environment for study participants and assessors. Due to two government-mandated, state-wide lockdowns, face-to-face-assessments were suspended from March 17 to June 24, 2020 and again from July 9 to October 20, 2020. Outside these dates, face-to-face assessments were offered where possible, but with physical distancing measures and hygiene procedures in place to minimise risk of viral transmission. Online telehealth-style assessments via a video conferencing platform were also developed and offered from June 12, 2020, so that families were able to take part while remaining in their own home. The latter involved an abbreviated assessment as certain measures could not be administered using telehealth (eg. movement and coordination items) (*Supplementary Table 1*). Families who took part in the telehealth-style assessment were invited to attend the hospital for a 3D craniofacial photo at a later date with the end to lockdown and when site visits became possible again.

### Participation rates (Table 1)

Of the 1342 eligible families, 802 completed the minimum data required for participation (60%) and neuropsychological assessment data are available for 696 children (52%). From commencement of the Covid-19 pandemic, 169 of the assessments were conducted in a telehealth format and another 73 in person with physical distancing in place. Following consent, we obtained externally assessed scores from the family’s private psychologist for nine children, which in two instances were complemented by a partial *AQUA at 6* assessment. Forty-one children who completed an assessment lived interstate, 23 of whom were visited by one of our assessors and 18 of whom completed a telehealth-style assessment. Another 14 children who lived overseas completed an assessment, three while visiting Melbourne and 10 via telehealth. (*data not shown*).

**Table 1.** *AQUA at 6* participation rates

|  | Eligible | Did not take<br>part | Questionnaire<br>completed | NP<br>assessment | Craniofacial<br>image | Brain<br>MRI | Buccal<br>swab |
| --- | --- | --- | --- | --- | --- | --- | --- |
| Invited | 1348 |  |  |  |  |  |  |
| Excluded | 6 |  |  |  |  |  |  |
| Unable to contact |  | 71 |  |  |  |  |  |
| No final response to any follow-up<sup>1</sup> |  | 161 |  |  |  |  |  |
| Opted out of AQUA at 6 |  | 308 |  |  |  |  |  |
| Partial questionnaire only |  |  | 12 |  |  |  |  |
| Questionnaire only<sup>2</sup> |  |  | 94 |  |  |  |  |
| Full neuropsychological assessment |  |  | 445<sup>3</sup> | 445 | 352 | 142 | 352 |
| Post-Covid neuropsychological assessment |  |  | 73 | 73 | 71 | 4 | 42 |
| Tele-neuropsychological assessment |  |  | 169<sup>4</sup> | 169 | 58 |  | 32 |
| Partial completion and/or external scores |  |  | 9 | 9 | 1 |  | 1 |
| **Total** | **1342 (100%)** | **540 (40%)** | **802 (60%)** | **696 (52%)** | **482 (36%)** | **146 (11%)** | **427 (32%)** |

<sup>1</sup> includes 49 mothers who agreed to the questionnaire but did not attempt to complete it.
<sup>2</sup> includes two questionnaires where child attended NP assessment, but unable to complete this.
<sup>3</sup> includes one partially completed questionnaire
<sup>4</sup> includes one partially completed questionnaire

Craniofacial photographs were obtained from 482 (36%) children. Participation rates in this aspect of the study were significantly impact by the two Covid-19-related lockdown periods where site visits were not possible.

Most of the brain MRIs were obtained prior to the Covid-19 pandemic, with an additional four children able to take part in the time following the lockdowns, resulting in 146 scans (out of a proposed 150) being available for analysis.

Buccal swabs were collected from 427 children, either while attending an in-person assessment or via home collection using a mailed swab kit.

540 eligible families did not take part in *AQUA at 6*: 308 opted out (23%); 71 for whom we had no current contact details (5%); and 161 who opted out passively either by not responding to any of our follow-ups or after initially expressing interest (12%).

Completion rates of previous post-birth study follow-ups in relation to *AQUA at 6* are presented in Table 2.

**Table 2.**
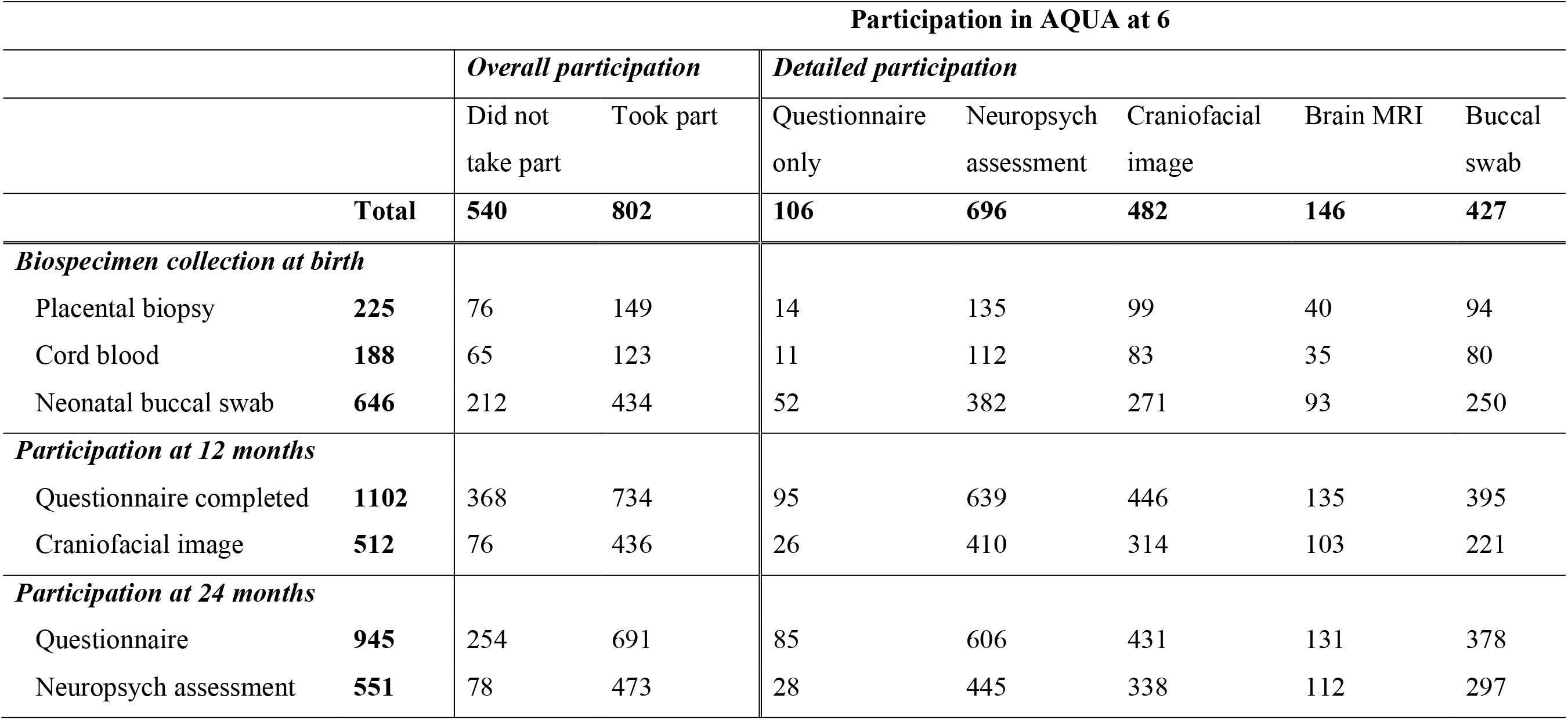
AQUA at 6 participation and availability of data from previous post birth follow-ups

### Exposure assessment

PAE patterns were assessed in the original AQUA study.[8] Complete data on drinking frequency, amount and type of alcoholic drink(s) on each occasion were collected for 1570 participants via three questionnaires administered in pregnancy.

#### Timing of exposure

Maternal alcohol consumption data were reported for five stages of pregnancy: (i) three months before pregnancy; (ii) trimester one pre-pregnancy aware; (iii) trimester one post-pregnancy aware; (iv) trimester two; and (v) trimester three. The mean (SD) gestational age at pregnancy recognition was 4.9 (1.5) weeks.[8]

#### Levels of exposure

Women were asked to use a pictorial drinks guide, listing common types and volumes of alcoholic drinks, to identify their ‘usual’ pattern of drinking, with provision for up to five types of alcoholic drink. For each beverage identified, they were asked how often they usually drank this type of alcohol and how many drinks they usually consumed on each occasion. Women were also asked if there were any ‘special occasions’ (or difficult times) when they consumed more alcohol than usual, the frequency of these occasions, the drink types, and the number of drinks per occasion. Estimates from ‘special occasions’ were combined with information from ‘usual’ alcohol consumption to calculate a maximum weekly intake.[8] The number and types of drink reported by women were firstly converted to standard drinks before calculating the amount of absolute alcohol in grams (gAA) consumed. One standard drink in Australia is equal to 10 gAA.

Alcohol abstinence throughout pregnancy (but not lifetime abstainer) was defined as the unexposed control group – no PAE.

Summarised exposure group data for the *AQUA at 6* eligible cohort (i.e. no PAE; PAE in trimester one only; PAE throughout gestation), and participation in the study’s core components are presented in Table 3. The PAE group distribution in the neuropsychological assessment and 3D craniofacial image data differed marginally from that in the eligible cohort, due to somewhat higher rates of participation in the ‘any PAE throughout pregnancy’ groups (41.8% and 43.4% respectively, compared to 37.6% in eligible cohort).

**Table 3.**
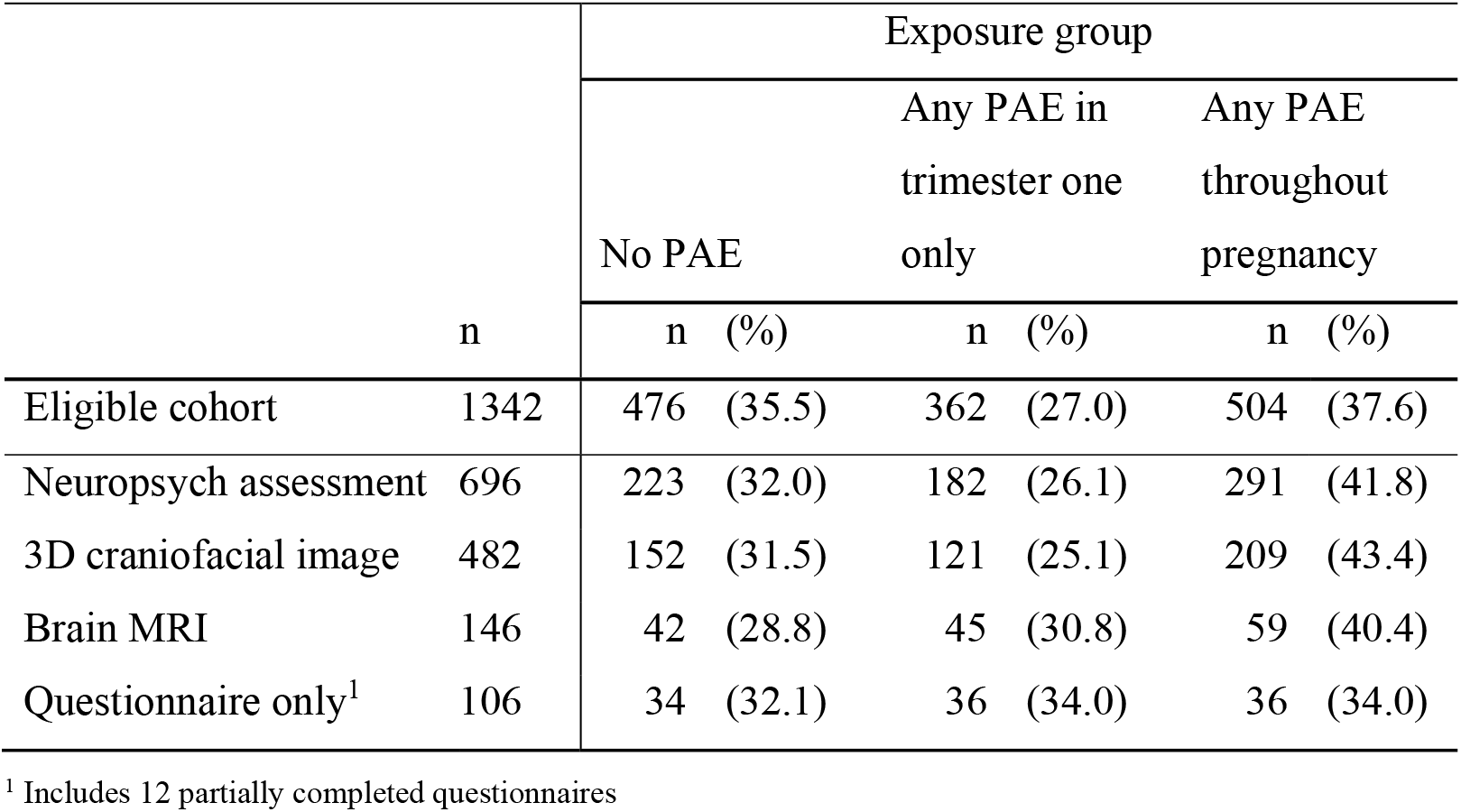
*AQUA at 6* participation by summarised prenatal alcohol exposure (PAE) group

In addition to this broad exposure classification, group-based trajectory modelling (GBTM) will be utilised as a data-driven method of classifying the temporal, continuous PAE data for all *AQUA at 6* analyses. GBTM can be used to objectively identify alcohol consumption trajectories arising directly from the source data without the need for pre-determined classification,[20] which has the potential to result in a more accurate and nuanced representation of the exposure to the fetus.

### Outcome measures

#### Neuropsychological Assessment

Children underwent a 3 to 4 hour neuropsychological assessment by trained psychologists blinded to PAE exposure and previous assessments (Table 4). The measures used were well-established and sensitive to early brain insult and were, based on measures identified as important in previous FASD research.[7, 21, 22] They include general intelligence (IQ), attention, executive function, memory and learning, language, and motor function.

**Table 4.** Neuropsychological Assessments

| Outcome domain | Scale/subtest |
| --- | --- |
| ***Psychologist assessed (direct assessment)*** |  |
| General Intelligence | Core subtests of the Wechsler Intelligence Scale for Children (WISC-V)[23] |
| Language | WISC-V Verbal Comprehension Index[23] |
| Academic functioning | Wechsler Individual Achievement Test (WIAT-III) subtests. Literacy is assessed using the word reading and spelling subtests, while mathematics is assessed using the numerical operations subtest[24] |
| Attention | Test of Everyday Attention for Children-Version 2 (TEA-Ch-2) subtests (*age* <8 *years*: Balloon Hunt, Barking, Sustained Attention to Response Task and Simple Reaction Time; *age* ≥8 *years*: Hector Cancellation, Vigil, Sustained Attention to Response Task and Simple Reaction Time)[25] |
| Working memory | Digits Recall (WISC-V), Blocks Recall and Backward Blocks Recall subtests of the Working Memory Test Battery for Children (WMTB-C)[25] |
| Cognitive flexibility | Contingency Naming Test (CNT trials 1-3)[26, 27] |
| Episodic memory | The California Verbal Learning Test – Children’s version (CVLT-C)[28] |
| Motor functioning | Movement Assessment Battery for Children (MABC2)[29] |
| ***Parent -report (indirect assessment)*** |  |
| Attention | ADHD Rating Scale 5[30] |
| Emotional & behavioural status | Strengths & Difficulties Questionnaire (SDQ)[31] |
| Executive function behaviours | Behavior Rating Inventory of Executive Function-Second Edition questionnaire (BRIEF-2)[32] |
| Autism symptoms | Social Communication Questionnaire (SCQ)[33] |
| Movement & coordination | Developmental Coordination Disorder Questionnaire (DCDQ07)[34] |

Neuropsychological assessments were complemented by information collected via a parent-report questionnaire using validated measures (Table 4).

#### Craniofacial imaging

Craniofacial imaging of the study child was undertaken by an experienced medical photographer using a 3dMD 7-pod system (3dMD corporation Atlanta GA, USA), which captures a full 360° image of the head (face and cranium). To ensure images were unobscured by hair, and to capture the shape of the neurocranium, a tight-fitting stocking was placed over the cranial vault. Images were captured in less than one second and available for review within three minutes. The photographer and craniofacial image analyst were blinded to the exposure.

To represent the entire surface of the cranium and face, a spatially-dense array of 69,587 points on an age-matched template face (pseudo landmarks), is automatically mapped onto each target image by a 3D surface registration algorithm. This warps the shape of the template into the shape of the target face, sampling each face at corresponding locations across the entire surface. Images are then able to be compared within and between groups using a robust generalised Procrustes analysis.[35]

#### Brain MRI

Brain imaging was undertaken at the Royal Children’s Hospital, Melbourne, using a 3 Tesla Siemens MAGNETOM Prisma scanner. The imaging sequences are listed in Table 5, with total time in the scanner around 45 minutes. Children in the MRI subgroup were scanned on the same day or within 2 weeks of their neuropsychological assessment. To ensure high compliance and quality images, children completed a preparatory session with a mock MRI scanner prior to their MRI appointment.

**Table 5.** MRI sequences

|  |  |
| --- | --- |
| 1 | T1- weighted multi-echo MP-RAGE images with 0.9 mm<sup>3</sup> isotropic voxels (IVs) and echo planar image-navigated prospective motion compensation |
| 2 | Multi-shell simultaneous multi-slice echo planar diffusion images (b= 750, 25 gradient directions; b=2000, 30 directions; and b=3000s/mm<sup>2</sup>, 45 directions) with 1.5mm<sup>3</sup> IVs and matching reverse phase encoding sequences |
| 3 | 3D T2-weighted turbo spin echo images with 0.9mm<sup>3</sup> IVs |
| 4 | Multiband, multi-echo gradient recalled echo planar resting state functional MRI images with 2.4 mm<sup>3</sup> IVs, with prospective acquisition correction and reverse phase encoding images |

Post-acquisition MR image analysis includes investigation of: (1) Regional brain volumes (66 cortical, 14 subcortical), and cortical morphology (thickness, curvature and sulcal depth) using FreeSurfer version 7;[36] (2) Volume and morphology of the corpus callosum, hippocampus, basal ganglia and cerebellum, regions hypothesised to be particularly important given previous FASD research;[1] (3) White matter microstructural organisation and maturity using (a) myelin mapping by applying the *T*1-*T*2 ratio,[37] (b) the Spherical Mean Technique, which can estimate diffusivity and neurite density from diffusion MR images without influence from crossing fibres,[38] and (c) Fixel Based Analyses, a fibre-based analysis of apparent fibre density; [39] (4) Whole-brain white matter tract analyses using tract-specific analyses,[40] as well as detailed examination of the corpus callosum, anterior-posterior fibre bundles, and corticospinal tracts using constrained spherical deconvolution tractography,[41] given their importance based on previous FASD research;[42] (5) Structural connectivity is examined using constrained spherical deconvolution-based white matter fiber tractography to find connections between FreeSurfer-derived brain regions. Using graph theory analyses, metrics such as global and local efficiency, small worldness, and rich club organisation will be produced to investigate the efficiency, integration or segregation of brain networks;[43] and (6) Functional connectivity analyses will also be done by applying Independent Component Analyses to resting state images, to find temporal correlations in spontaneous blood oxygen level-dependent signal between brain regions.[44]

#### Australian Early Developmental Census (AEDC)

Consent was sought to link *AQUA at 6* children who attended their first year of school in 2018 to the AEDC,[45] and 93% of mothers with an eligible child consented to this data linkage. The AEDC is undertaken every three years using the Australian version of the Canadian Early Development Instrument,[46] with the most recent year being 2018. The instrument consists of 100 questions and is completed by teachers on the basis of at least one month’s knowledge of the child. It covers the five domains of physical, social, emotional, language and cognitive development, as well as data on special needs. Children falling below the 10th percentile in any domain are considered developmentally ‘vulnerable’ in that area, children falling between the 10th and 25th percentile are considered developmentally ‘at risk’, and all other children are considered to be ‘on track’. Approval for linkage to relevant data items at an individual (micro) level will be obtained from the AEDC custodians and the linkage will be conducted independently by an authorised Data Linkage Agency.

### Confounders and modifiers

During the original 2011-2014 AQUA study, extensive data were collected on factors that may confound or modify the relationship between PAE and child outcomes. These included maternal obstetric history and complications, maternal nutrition and supplementation, breastfeeding, maternal and paternal drug use, maternal mental health, education and other socio-demographics, family relationships, and parenting [19]. Updated relevant information on demographic and socio-environmental factors was collected from the child’s primary caregiver (Table 6).

**Table 6.**
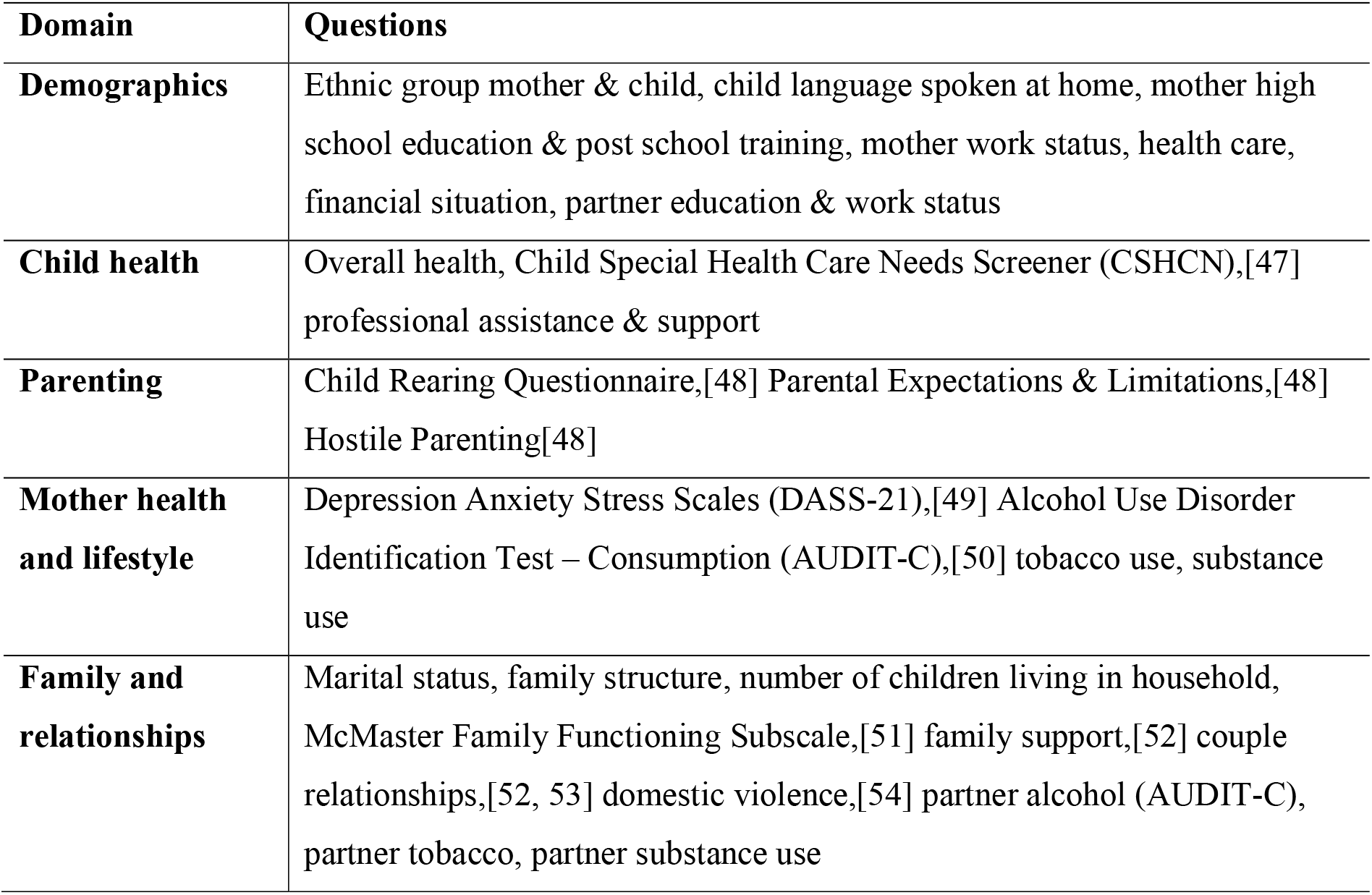
Demographic and socio-environmental factors collected by parent-report

| Domain | Questions |
| --- | --- |
| **Demographics** | Ethnic group mother & child, child language spoken at home, mother high school education & post school training, mother work status, health care, financial situation, partner education & work status |
| **Child health** | Overall health, Child Special Health Care Needs Screener (CSHCN),[47] professional assistance & support |
| **Parenting** | Child Rearing Questionnaire,[48] Parental Expectations & Limitations,[48] Hostile Parenting[48] |
| **Mother health and lifestyle** | Depression Anxiety Stress Scales (DASS-21),[49] Alcohol Use Disorder Identification Test – Consumption (AUDIT-C),[50] tobacco use, substance use |
| **Family and relationships** | Marital status, family structure, number of children living in household, McMaster Family Functioning Subscale,[51] family support,[52] couple relationships,[52, 53] domestic violence,[54] partner alcohol (AUDIT-C), partner tobacco, partner substance use |

### Biospecimens

A comprehensive biobank of maternal and child DNA from the AQUA study exists, comprising extracts from placental biopsies, cord blood mononuclear cells and maternal and neonatal buccal swabs to investigate how genetic, epigenetic and environmental factors interact with PAE to explain individual variation in child outcomes.[19] This biobank was extended for future collaborative investigations in this area through the collection of 427 child buccal swabs in 6-8-year-olds (Table 1).

## FINDINGS TO DATE

In the original cohort there were 1570 mother-child pairs, of whom 59% of mothers reported drinking alcohol during pregnancy and 19% reported at least one episode of binge drinking prior to pregnancy recognition.[8] The study found an association between low to moderate PAE and craniofacial shape in the children aged 12 months, with differences concentrated around the nose, eyes, and mouth.[55] This has potential clinical implications given that development of the face parallels, and is controlled by, the brain. However, at two years of age, no adverse association was detected between child neurodevelopment and low to moderate PAE using the Bayley Scales of Infant and Toddler Development (Bayley-III).[56] Given that measures of early development are only moderately predictive of school-aged outcomes,[57] and do not reliably assess higher-order cognitive and motor functions,[58, 59] this school-aged follow-up of this cohort is essential to determine any long-term effects of low to moderate PAE and binge episodes.

## POWER CALCULATIONS

We have many functional outcomes, an important one being full-scale IQ (SD = 15). With a 52% participation rate and with a two-sided 0.05 significance level, we will have 80% power to detect a small but clinically meaningful effect size (Cohen’s f=0.12), equating to a mean difference of 3.6 IQ points between the three major PAE groups. In terms of the MRI data, an important measure of interest is brain volume. Assuming a mean intracranial volume of around 1350 cubic cm (cc) (SD = 120cc) [60], which is based on typically developing seven-year-old children in Melbourne, with a sample size of 146 we will have 80% power to detect a difference of 64cc in total brain volume between groups (medium effect size f=0.26). Traditional power calculations are not possible for craniofacial analysis, where the outcome measure is many thousands of point coordinates. In our one-year analysis we detected differences (p<0.05) using a control group of 89 and PAE groups of approximately 40 images,[55] which gives us confidence that our proposed analyses will be sufficiently powered.

## STRENGTHS AND LIMITATIONS

This study has several unique features that will enable the relationship between PAE and neurodevelopment to be addressed rigorously. Firstly, AQUA has very detailed assessments of drinking patterns in the peri-conceptional period and during pregnancy, including timing, frequency and quantity of alcohol consumption. Secondly, AQUA is a large representative cohort of pregnant women from the community, recruited for the specific purpose of assessing common drinking patterns. Previous studies have tended to focus on high-risk groups of substance users, risking selection bias, or take advantage of crude measures collected during large epidemiological studies.[15, 61, 62] Thirdly, at each phase of data collection we have gathered information relating to important modifiers and confounders including nutrition, mental health, physical health, and socio-economic status. Adjustment for these factors is essential given their relationship to both drinking behaviour and child development. Finally, AQUA uses sensitive measures targeting specific areas of neurodevelopment informed by FASD research including longitudinal analysis of craniofacial shape, brain structure and function, and neuropsychological functioning. Of importance, our investigations include whether early craniofacial changes are predictive of later neuropsychological impairments, and whether PAE is associated with a common pattern of neural abnormalities demonstrated on MRI.

A limitation of any study measuring PAE is that there are currently no validated objective measures to quantify low to moderate exposure,[63] and researchers depend on accurate maternal recall and reporting. In order to maximise accuracy of reporting in the AQUA study, we involved pregnant women in the development of the alcohol consumption questions to be used in the AQUA study.[64] This work indicated that women who attend general antenatal care would answer as truthfully as possible, due to their vested interest in understanding what may be considered normal, non-risky pregnancy drinking habits. The opportunity to report heavy or binge drinking on ‘special occasions’ yielded important information on early gestation exposures, information that might not have been reported in more general questioning.[8]

The validity of some covariates such as maternal lifestyle and family relationships may also be subject to reporting bias due to a desire to provide socially acceptable responses. Findings will need to be interpreted in the context of existing literature on the causal relationships between such variables and child neurodevelopment.

Finally, in instances when direct neuropsychological assessment of the child was not possible, we depended on indirect measures (e.g. maternal report) to determine developmental progress, which is subjective and may introduce informant bias.

In summary, a significant proportion of pregnant women do not adhere to health policy guidelines and drink some alcohol, potentially putting thousands of children at risk for life-long neurodevelopmental impairments. No safe level of alcohol consumption in pregnancy has been established, and women’s drinking behaviour in part reflects the lack of evidence to support health professional advice that women who are pregnant should not drink alcohol. Findings from this study will have an impact from a preventative health perspective, providing strong evidence on the consequences of low to moderate and binge-level prenatal alcohol exposure, strengthening the messages provided to the public through education and health promotion campaigns.

## Data Availability

The datasets generated and/or analysed during the current study will not be publicly available as the study is ongoing. The raw data supporting the conclusions of future manuscripts will be made available to the journal's supplementary material by the authors, if requested. AQUA at 6 study families have the option to consent for their data to be used in future related and ethically approved projects. Following study completion, data will be available from the corresponding author upon reasonable request to such projects.

## COLLABORATIONS

As with our 12-month follow-up, we are collaborating with two experts in 3D morphometric analysis of image data: Drs Peter Claes and Harold Matthews from KU Leuven in Belgium. The collaboration aims to develop new approaches to undertake our craniofacial analysis and to interpret these results.

Other collaborations to-date have arisen from our interest in epigenetics. Specifically, the association between PAE and DNA methylation and its role as a mediator of neurodevelopment and FASD. We are contributing data to the Pregnancy And Childhood Epigenetics (PACE) consortium as part of their meta-analysis project studying early life environmental impacts on human disease using epigenetics. The consortium is based at the US National Institute of Environmental Health Sciences and includes researchers from around the world. (https://www.niehs.nih.gov/research/atniehs/labs/epi/pi/genetics/pace/index.cfm).

We are also collaborating with the lab of Professor Michael Kobor, Centre for Molecular Medicine and Therapeutics, BC Children’s Hospital Research Institute, The University of British Columbia. The Kobor lab recently developed a pediatric epigenetic clock (PedBE) using buccal epithelial swabs (https://github.com/kobor-lab/Public-Scripts/). The collaboration will generate epigenetic and genotypic data from our child buccal DNA to contribute to their project investigating the extent to which the PedBE clock informs on child development across diverse populations and sex. Another collaboration in this area of study is with a team at the Telethon Institute, Western Australia, led by Dr David Martino. The AQUA study is contributing EWAS data from buccal epithelial swabs for this project, which aims to identify DNA methylation biomarkers of PAE in a controlled murine experiment, with replication in existing methylation data sets from human infants with well characterized PAE exposure patters and children diagnosed with FASD. (https://www.telethonkids.org.au/contact-us/our-people/m/david-martino/).

The AQUA study welcomes new collaborations with other investigators and have actively engaged in collaborative data-sharing projects. Interested investigators should contact the Project Manager Evi Muggli to obtain additional information about the study and referral to the appropriate Chief investigators for the discussion of collaborative opportunities.

The AQUA study has obtained participant consent to have their data included in other ethically approved studies in related areas of research.

## FURTHER DETAILS

### Data management

All study data are collected and managed using REDCap electronic data capture tools hosted at The Murdoch Children’s Research Institute in Melbourne, Australia.[65, 66] REDCap (Research Electronic Data Capture) is a secure, web-based software platform designed to support data capture for research studies, providing 1) an intuitive interface for validated data capture; 2) audit trails for tracking data manipulation and export procedures; 3) automated export procedures for seamless data downloads to common statistical packages; and 4) procedures for data integration and interoperability with external sources. REDCap is also used to facilitate tracking and scheduling of all communication with participants. Electronic raw and derived data, including longitudinal data from the first phase of the AQUA study, will be stored on a restricted server and curated by the Project Manager (EM).

### Ethics approval and consent to participate

All study procedures have been approved by the Human Research Ethics Committee of the Royal Children’s Hospital, Melbourne, Australia according to the National Statement on Ethical Conduct in Human Research (2007) issued by the National Health and Medical Research Council of Australia (approval #38025). Project staff obtained written informed consent from the child’s guardian, i.e. the mother in most cases. All personal information of potential and enrolled participants was collected only for research purpose and will be kept in strict confidentiality by the investigators and project staff. Personal information is stored separately from other research data and will be linked using the family’s study ID which was assigned at enrolment of the pregnant mother in the first phase of the AQUA study. Hard copy materials are kept in locked compartments and electronic records are stored with password encryption. All hard-copy and electronic data are stored until child participants are 25 years of age or for fifteen years after the study has been completed, whichever is later.

### Availability of data and materials

The datasets generated and/or analysed during the current study will not be publicly available as the study is ongoing. The raw data supporting the conclusions of future manuscripts will be made available to the journal’s supplementary material by the authors, if requested. *AQUA at 6* study families have the option to consent for their data to be used in future related and ethically approved projects. Following study completion, data will be available from the corresponding author upon reasonable request to such projects.

### Patient and public involvement

Patients and/or the public were not involved in the design, or conduct, or reporting, or dissemination plans of this research.

## Acknowledgements

The authors would like to thank all AQUA study families who generously gave their time to participate in this study to date. We are extremely grateful to our dedicated staff and students whose enthusiasm never failed despite the Covid-19 pandemic and who recruited and assisted the families through their study participation, undertook the neuropsychological assessments and MRI training and provided feedback to families; Sophie Gibson, Pip Pyman, Garance Delagneau and Drs Stephanie Malarbi, Ngoc Nguyen and Siobhan Seward-Swann. We also thank Stephanie, Ngoc and Siobhan for their invaluable contribution to the Covid-19 related adaptations of the neuropsychological assessment protocols.

## Authors’ contributions

All authors were involved in the conception and in the design of the study and have made substantial contributions to the planning of the study. EM drafted the manuscript. All authors critically revised manuscript drafts and agreed with the final version of the manuscript.

---

**Supplementary Table 1.**
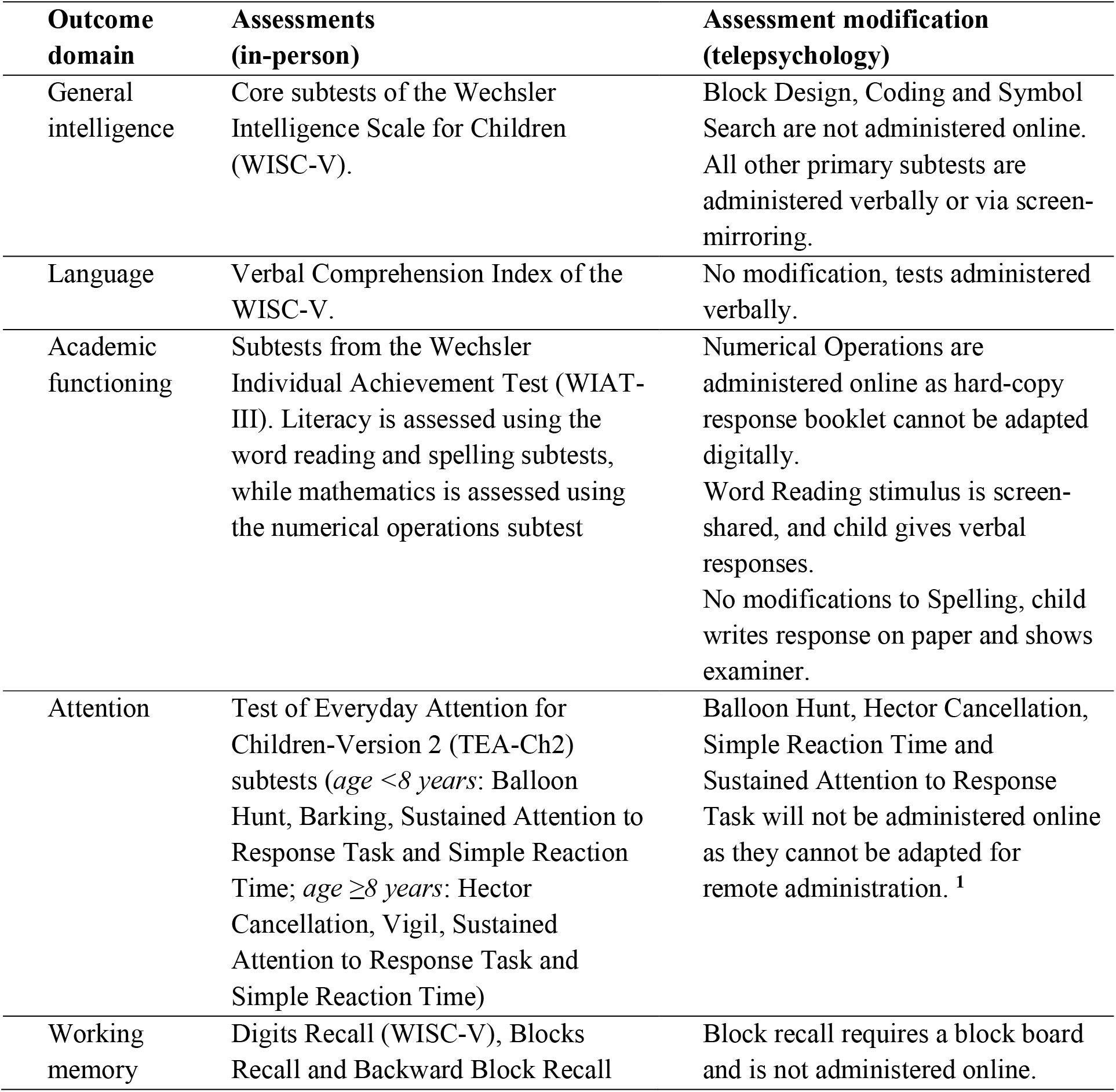
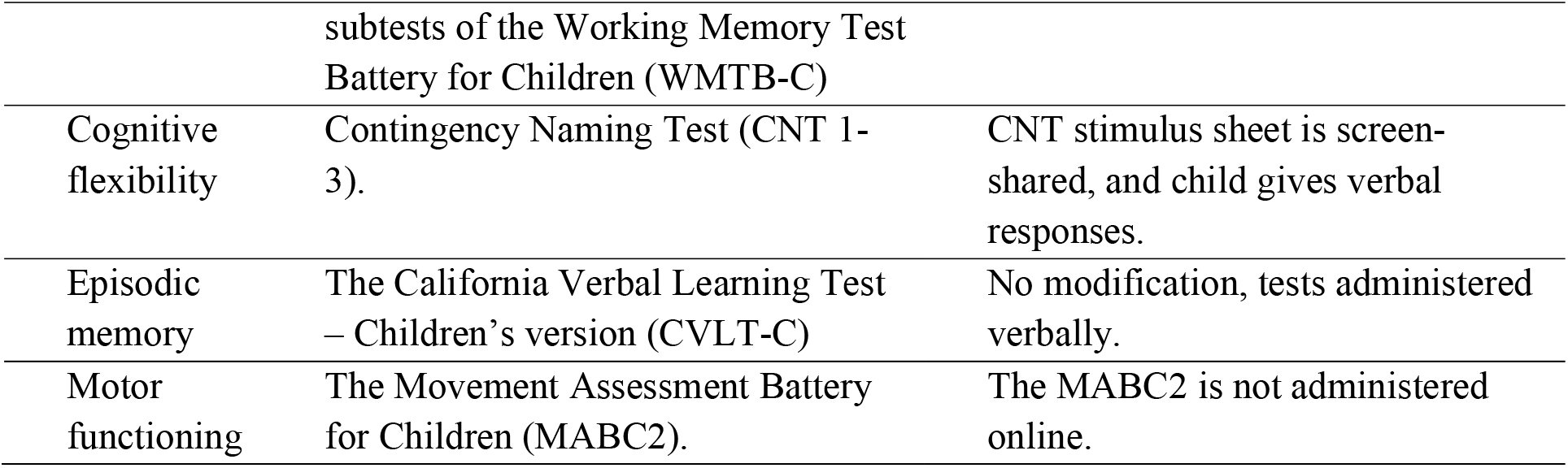
Telepsychology modifications to the in-person assessment protocol

